# Digital Nursing Quality Management: A Concept Analysis

**DOI:** 10.1101/2023.07.16.23292725

**Authors:** Lin Han, Xinrui Bai, Hongyan Zhang, Ning An

**Affiliations:** Evidence-based Nursing Center, School of Nursing, Lanzhou University, Lanzhou City, Gansu Province, China; Department of Nursing, Gansu Provincial Hospital, Lanzhou City, Gansu Province, China; School of Computer Science and Information Engineering, Hefei University of Technology

**Keywords:** Concept analysis, Digital health, Digital Nursing Quality Management, Health information technology, Nursing Staff, Hospital, Nursing Administration Research

## Abstract

**Background:** The rapid advancements in digital technology and its increasing integration into healthcare create an opportunity to leverage these innovations for enhancing nursing quality management.

**Objective:** This paper aims to report the results of a concept analysis of digital nursing quality management.

**Methods:** Rodgers’ Evolutionary Method of concept analysis was used to examine the attributes, antecedents, consequences, and related concepts of digital nursing quality management.

**Results:** Digital Nursing Quality Management is a strategic management approach that systematically utilizes evolving digital technology to pursue optimal nursing quality, guided by the timeless principle of humanistic care. The defining attributes are human-machine collaborativeness, interactive empowerment, and digital ecosystem.

**Conclusions:** Digital nursing quality management is not a mere amalgamation of separate components but a dynamic ecosystem shaped by individual, organizational, and technological factors.

## Introduction

Digital technologies have transformed various sectors, and healthcare is no exception (Navarro-Martínez et al., 2023). Integrating digital technologies into nursing practice, from electronic health records (EHRs) to telehealth and mobile health applications, has opened new avenues to enhance patient care, improve health outcomes, and optimize nursing workflows (Philpot et al., 2023). One emerging area of focus in this digital transformation is Nursing Quality Management, a fundamental aspect of nursing practice to ensure high-quality care and patient safety. As digital technologies become increasingly embedded in nursing practice, it is imperative to understand and harness their potential in advancing Nursing Quality Management. This shift has led to the conceptual evolution of “Digital Nursing Quality Management.”

Historically, Nursing Quality Management has focused on implementing standards and guidelines, promoting best practices, and continuously monitoring and improving the quality of nursing care to enhance patient outcomes (Ebneter et al., 2022). While these traditional approaches remain essential, incorporating digital technologies provides novel opportunities for more efficient, data-driven, and personalized quality management (Papachristou et al., 2023). Digital Nursing Quality Management encapsulates these opportunities, bridging the intersection of digital health and quality management in nursing practice.

Despite its growing relevance, the concept of Digital Nursing Quality Management is relatively underexplored in the nursing literature. A clear and comprehensive understanding of this concept, including its attributes, antecedents, consequences, and application in healthcare settings, is crucial for researchers, nursing professionals, and healthcare organizations. Defining the elements and characteristics of this concept will guide the effective integration of digital technologies into nursing quality management, inform the development of educational and training programs for nurses, and shape future research and policy in this area.

Therefore, this paper conducted a concept analysis of Digital Nursing Quality Management. We systematically review the existing literature and use Rodgers’ evolutionary concept analysis method to explore and define the concept, identify its core attributes and related concepts, and understand its application in healthcare. Through this analysis, we aim to contribute to the ongoing discourse around digital health in nursing practice and provide a foundation for further research and practice developments in Digital Nursing Quality Management.

## Methods

Rodgers’ evolutionary concept analysis method is well suited to this activity cause of its systematic method and applicability when exploring fluid or changing concepts (Tofthagen and Fagerstrøm, 2010). This approach involves six phases (Rodgers, 2000): (a) Concept identification; (b) Determining the scope of data collection; (c) Collecting data on the concept regarding attributes and context; (d) Analysing the data; (e) Providing an example of the concept in use, if appropriate; and (f) Identifying implications, hypotheses, and future development needed. According to Rodgers (Risjord, 2009), concepts evolve with the advancement of theories and practices. Thus, the defining characteristics of a concept identified using this method somewhat reflect the historical context.

### Data sources

A comprehensive electronic literature search of PubMed, Web of Science, CINAHL, Cochrane Library, China National Knowledge Infrastructure, Wanfang Data, SinoMed Database, and VIP Database was conducted from January 1, 2000, to June 10, 2023. The keywords employed were digital nursing, digital care, quality of care, quality of nursing, and quality of nursing management, and the search terms were used individually or in combination with each other. The study noted that since 2000, “digital health” and “digital” as a term have appeared frequently in research. Interest in digital health has grown exponentially, and articles published before 2000 were excluded to ensure the quality of the included literature (Brommeyer et al., 2023).

### Literature analysis

The Digital Nursing Quality Management concept was examined using Rodgers’ Evolutionary concept analysis. A trained graduate student and experienced professor participated in the analysis. Before the literature search, researchers developed search strategies and plans for analysis. To ensure high-quality analysis, Initially, the full paper was read, and the extracted data were classified into an Excel spreadsheet and categorized the defining characteristics, antecedents, and consequences. Then, the same or similar content was combined to compare and classify all extracted content until the concept features became clearer.

## Findings

### Search Results

The initial exploration identified 3379 papers; the duplicates (n = 299) were excluded. After reviewing the titles and abstracts of the remaining articles, 2764 literatures were removed, and the full texts of 316 studies were evaluated. Overall, 88 publications were included in this study, including 57 English articles and 31 Chinese academic journal articles. The search process followed the Preferred Reporting Items for Systematic Reviews and Meta-Analysis (PRISMA) search strategy guidelines presented in Fig. 1.

**Figure 1.**
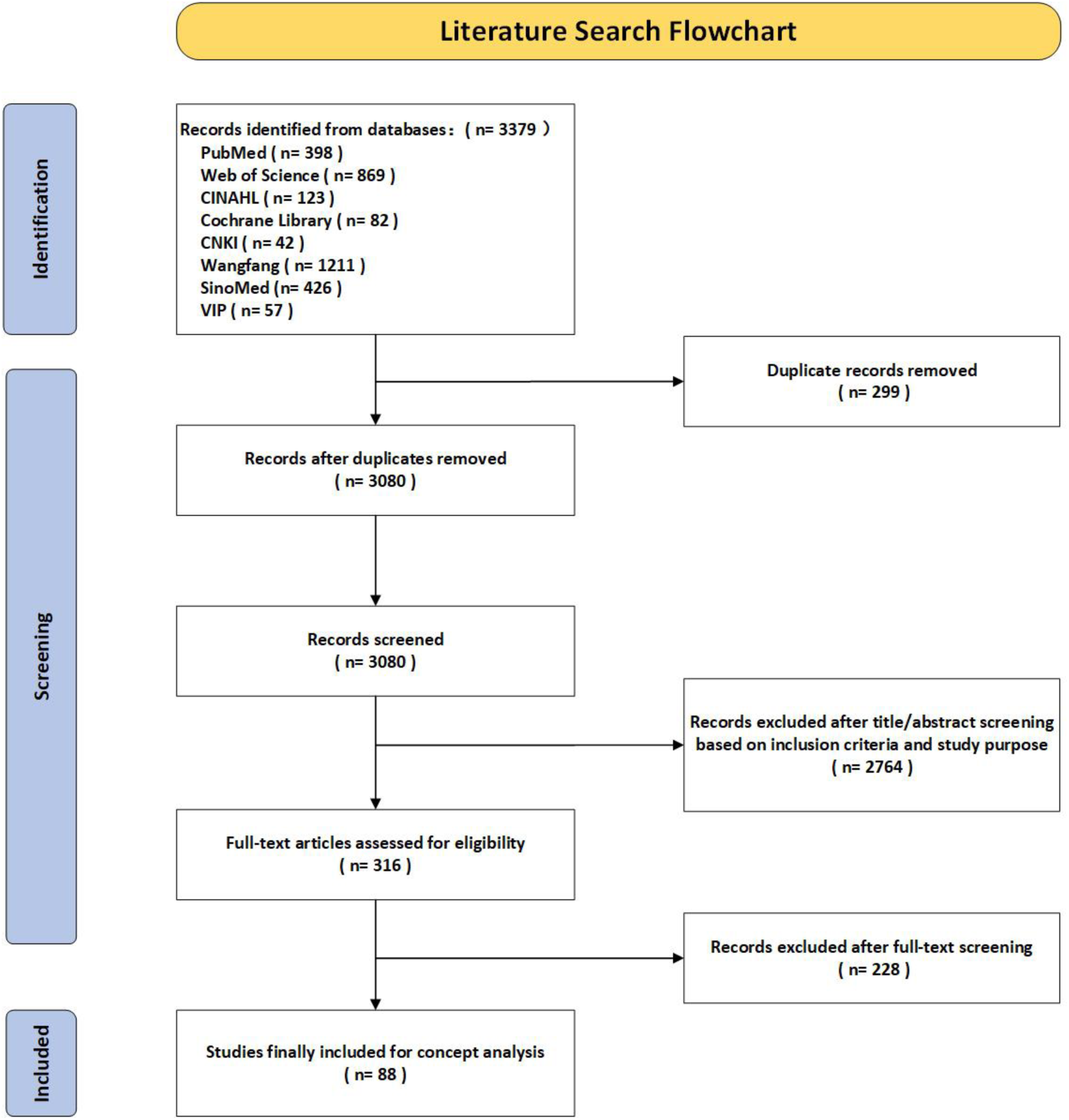
Search strategy PRISMA diagram.

Table 1 highlights 23 of the 88 articles because they each cover the concept’s attributes, antecedents, and consequences and are particularly informative. Although the other 65 articles do not comprehensively refer to all concept elements, they provide equally essential clues in identifying a particular structure of the conceptual framework and contribute to the development of the conceptual model. Next, a realistic model case was retrieved to demonstrate the concept. Finally, a proposed definition of digital nursing quality management was established. A visual depiction of the components of DNQM derived from the concept analysis is presented in Fig. 2. The following subsections discuss the findings of this paper.

**Table 1.** Explanation of the concept of digital nursing quality management in 23 core articles.

| Authors, Date | Concept-related description | Attributes | Antecedents | Consequences |
| --- | --- | --- | --- | --- |
| (Bauer and Brown, 2001) | The digital transformation of healthcare | • Digital Ecosystem<br>• Interactive Empowerment | • Computer-supported telemedicine | • Patients get timely care<br>• Improvement of personnel expertise and skills |
| (Jaramillo, 2001) | A systematic approach to data management | • Digital Ecosystem<br>• Human-machine collaboration | • Management innovation<br>• Disease management data warehouse | • Facilitating the processes of implementing algorithms, analysing data, and reporting on information on clinical, financial, and operational outcome measures |
| (Kalichman et al., 2002) | Changes brought about by interactive computer-based health information technologies | • Human-machine collaboration<br>• Interactive Empowerment | • Individual's knowledge, motivation, and skills<br>• Health information systems | • Motivating patients to engage in their healthcare and improve their health behaviours |
| (Roeder, 2009) | Electronic integrated system for extracting data to monitor quality management issues | • Digital Ecosystem<br>• Human-machine collaboration<br>• Interactive Empowerment | • Technical implementation team<br>• Agency conducts digital skills training<br>• Life cycle of electronic systems and information storage capacity | • Freeing nurses from written records<br>• Reducing redundant tests and medical errors and improving hand-off communications |
| (Litwin et al., 2018) | A clinical decision-making tool to enhance service delivery and improve the quality of care | • Digital Ecosystem<br>• Human-machine collaboration | • Technical coordinator<br>• Facilities support from the organization | • Increasing visibility of labour management data<br>• Iterative user feedback |
| (Oberg et al., 2018) | Using digital e-health systems to support patient management | • Human-machine collaboration<br>• Interactive Empowerment | • Shortage of skilled staff<br>• Local authorities and institutions work to promote digital care | • Patient self-management<br>• Conducive work environment<br>• Lack of personalized care |

**Table 1** (Continued).
| Authors, Date | Concept-related description | • Attributes | • Antecedents | • Consequences |
| --- | --- | --- | --- | --- |
| (Au et al., 2019) | Visual analysis and management of patients' conditions | • Digital Ecosystem<br>• Human-machine collaboration<br>• Interactive Empowerment | • Digital wound care management solutions<br>• Coordinator<br>• Inter-organizational comparison of quality ratings | • Facilitates tracking of disease<br>• Less workload<br>• Rights protection |
| (Dugstad et al., 2019) | A novel set of behaviours, routines, and ways of working that are directed at improving health outcomes, administrative efficiency, cost-effectiveness, or users' experience and that are implemented by planned and coordinated actions | • Digital Ecosystem<br>• Human-machine collaboration | • Digital monitoring technology systems<br>• Technology absorption capacity of the organization<br>• Multi-sectoral collaboration | • Identification and redistribution of roles and responsibilities<br>• Potential cost reduction<br>• Nurturing the development of mutual trust and a constructive dialogue |
| (Egbert et al., 2019) | A new approach to integrating the competency areas of nursing informatics more closely with those of nursing research, quality assurance, and healthcare management | • Digital Ecosystem<br>• Human-machine collaboration | • Health IT applications<br>• Connectivity between healthcare providers<br>• Management-related core competency | • Improving patient outcomes and the quality and efficiency of care processes |
| (Hernandez, 2019) | Machine simulation of the role of a nurse | • Human-machine collaboration<br>• Interactive Empowerment | • Chatbot care<br>• The agency framework of organizational communication | • Bridging the gap between the demand and supply of human healthcare providers |
| (Thomsen et al., 2019) | Using mobile processes to improve the quality of care | • Digital Ecosystem<br>• Human-machine collaboration<br>• Interactive Empowerment | • Mobile applications<br>• Clinical nursing practice demands | • Improving the knowledge, skills, and confidence of health workers<br>• Improving patient acceptance of the technologies |

**Table 1**(Continued).
| Authors, Date | Concept-related description | Attributes | Antecedents | Consequences |
| --- | --- | --- | --- | --- |
| (Sun and Mo, 2019) | A product of the close integration of information technology and medical activities | • Digital Ecosystem<br>• Interactive Empowerment | • Nursing quality management system | • Standardisation of nursing indicators and quality of care checks across the hospital |
| (Hong et al., 2020) | Healthcare providers and organizations provide information and services to patients and caregivers through digital tools | • Digital Ecosystem<br>• Human-machine collaboration<br>• Interactive Empowerment | • Penetration of digital tools<br>• Audience acceptance of technologies | • Effective and efficient management<br>• Better relationships between the patients and providers<br>• Improving access to healthcare services |
| (Melchiorre et al., 2020) | The application of innovative Information and Communication Technologies (ICT) in nursing delivery | • Digital Ecosystem | • Availability of ICT infrastructures, services, and skills | • Improving the quality of available data<br>• Improving the safety and quality of patient care |
| (Whitmore et al., 2020) | Using digital health interventions to help and manage the diseases of patients | • Digital Ecosystem<br>• Human-machine collaboration<br>• Interactive Empowerment | • Clinical Practice Changes | • Management simplification<br>• Enhancing quality of care |
| (Ceyhan et al., 2021) | Utilizing information systems in the nursing industry to improve nursing quality | • Human-machine collaboration | • Increased use of information technology<br>• Shortage of technical nurses | • Creating a common language for nursing plans |
| (Wei et al., 2021) | A data-driven digital platform for quality control management | • Digital Ecosystem<br>• Human-machine collaboration | • Multi-sectoral collaboration<br>• Technology support | • Improving efficiency and quality<br>• Refinement of nursing quality management |
| (Back et al., 2022) | Improving care through digital health procedures | • Digital Ecosystem<br>• Human-machine collaboration | • Digital doctor-patient communication quickly grew in importance<br>• Digitally integrated care structure | • Cost reduction<br>• Changes in mindset, process, and structure |

**Table 1**(Continued).
| Authors, Date | Concept-related description | Attributes | Antecedents | Consequences |
| --- | --- | --- | --- | --- |
| (Chen et al., 2022) | A mobile app to help nurses manage medical equipment and substances | • Digital Ecosystem<br>• Human-machine collaboration | • The actual need to simplify nursing work | • Improving nurses' work satisfaction, quality of care, and workload reduction |
| (Lee and Chang, 2022) | Enhancing patient health outcomes by improving nurses' digital competencies | • Human-machine collaboration<br>• Interactive Empowerment | • Development of information technology<br>• Patient demand for high-quality care | • Improving critical thinking for managers<br>• Improving the quality of care |
| (Sly et al., 2022) | Utilizing data-visibility solutions to promote remote and targeted reviews of patients | • Digital Ecosystem<br>• Human-machine collaboration<br>• Interactive Empowerment | • Digital clinical decision support systems (CDSS)<br>• Medical structure's willingness to optimize its structure | • Supporting clinical decision-making<br>• Improving the effectiveness of patient safety and management |
| (Bail et al., 2023) | A novel point--of--care digital health management | • Digital Ecosystem<br>• Human-machine collaboration<br>• Interactive Empowerment | • Digital health information systems | • Improving satisfaction of staff and residents<br>• Improving management decisions |
| (Hoyle, 2023) | Digital solutions to support patient safety should be actively promoted and implemented | • Digital Ecosystem<br>• Human-machine collaboration | • Software performance<br>• Organizational support | • Freeing nurses from written records<br>• Increased risk identification capabilities |
| (Laine et al., 2023) | Technical tools to support quality of care management | • Human-machine collaboration | • Instrument performance | • Reducing cognitive burden and errors<br>• Improving care processes, outcomes, and situational awareness |

**Figure 2.**
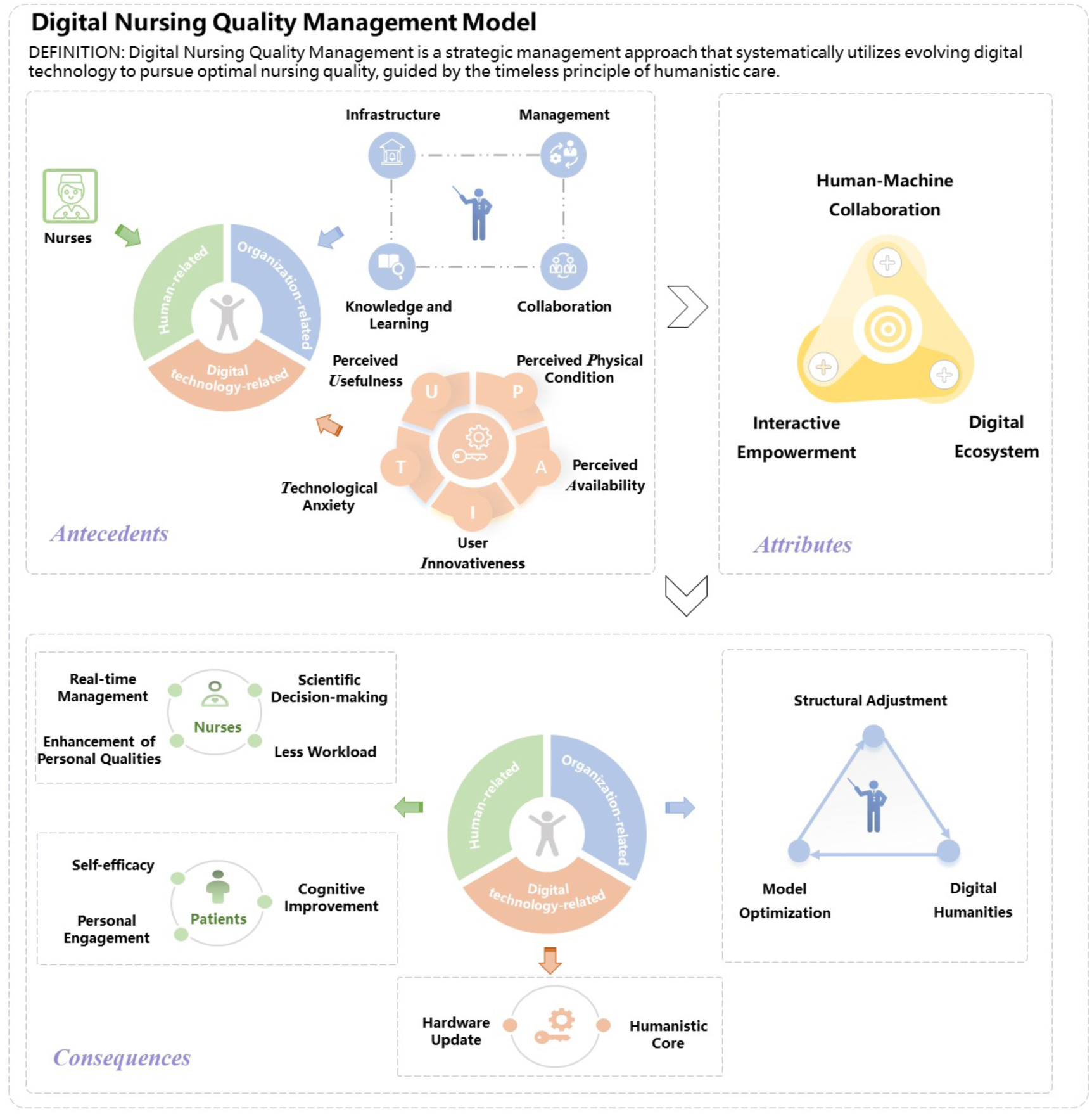
Model of Digital Nursing Quality Management.

### Attributes of digital nursing quality management

Attributes are the characteristics or phenomena of a concept (Rodgers et al., 2018). Specifying the critical attributes of a concept is an essential step in its clarification and refinement, which results in a deep understanding of the concept and its differentiation from other concepts (Rodgers, 2000). The attributes of digital nursing quality management are human-machine collaboration, interactive empowerment, and digital ecosystem.

#### Human-machine collaboration

Human-machine collaboration is a combination of human intelligence and machine capabilities that works best when actors complement each other’s strengths (Inga et al., 2023). It goes one level further than human-machine interaction, incorporating collaboration between nurse-machine and patient-machine, and is crucial in digital nursing quality management.

##### (1) Nurse-Machine Collaboration

Collaboration indicates that both nurses and technologies contribute “labour” to patient-centred care practice (Garrett and Klein, 2008, Roeder, 2009), the former “tangible” and the latter “intangible.” The incorporation of technology or programs enables nurses to be less constrained by changes in a patient’s outwardly visible condition, to more fully track the source of the cause, and to clarify the therapies that can be done in the first instance(Choi and Tak, 2022). Digital technologies like artificial intelligence, electronic health records, telehealth platforms, and decision-support systems help nurses streamline workflows, provide access to real-time patient data, assist in clinical decision-making, and improve the quality of care (Arakawa et al., 2018, Kalichman et al., 2002).

##### (2) Patient-Machine Collaboration

“Use” is a crucial sign of patient-machine collaboration, and patients’ “perceived usefulness” and “perceived ease of use” of digital technologies impact the extent and efficacy of human-machine collaboration via patients’ desire to use. When patient-facing technologies like mobile health applications, wearable devices, and personal health records provide valuable data for nursing assessments and interventions, as well as improve patient satisfaction and health outcomes, the degree of patient collaboration with digital technologies evolves (Parreira et al., 2018). Additionally, factors like the accessibility and user-friendliness of technologies, patients’ digital health literacy, and technology integration into care plans contribute to this collaboration (Dohse, 2021).

In the following sections, we will elaborate on the other critical attributes of digital nursing quality management – interactive empowerment and digital ecosystem – and explore their implications for nursing practice and research.

#### Interactive empowerment

The era of digital health has witnessed a profound shift in the dynamics of healthcare, with a growing emphasis on empowering healthcare professionals and patients to participate in care processes actively. In the context of Digital Nursing Quality Management, this empowerment takes an interactive form.

##### (1) Empowerment of Nurses

The provision of applications, platforms, and resources enables more nurses to put their ideas into practice and improve patient care (Gross et al., 2021). For example, advanced analytics from EHRs aid in predicting patient outcomes and evidence-based clinical decisions (Roeder, 2009). Digital communication platforms enhance patient engagement and enable nurses to contribute to designing and implementing digital tools, ensuring that they meet their needs and improve the quality of care (Litwin et al., 2018).

##### (2) Empowerment of Patients

Digital technologies empower patients by enabling active participation in their care through mobile health applications, telehealth services, and wearable devices (Bally et al., 2023, Lober and Flowers, 2011). This allows them to monitor health conditions, access information, and communicate effectively with healthcare providers, promoting the experience of self-efficacy and better coping (Gross et al., 2021).

##### (3) Empowerment of Organizations

Digital technology fundamentally empowers healthcare organizations in multiple ways. Firstly, digitalization provides managers with the possibilities for “seeing”, “knowing”, and “governing” actions, which aids in streamlining internal processes, promoting effective collaboration, facilitating the sharing of information and decisions, and even creating a digital culture that encourages innovation and continuous learning (Balta et al., 2021, Cherbib et al., 2021). Secondly, digitized data collection paired with real-time analytics offers significant support for quality monitoring and improvement initiatives at the regional and national levels. Lastly, one of the more impactful potentialities of this empowerment is those platform-wide data-driven insights make it clearer where specific organizations can be remediated, providing the right direction for improvement and thus improving the quality of care delivery across the network (Cannavacciuolo et al., 2023). This broad-based interactive empowerment fosters a more collaborative and efficient healthcare system, enabling healthcare organizations to respond more effectively to evolving patient needs and healthcare standards (Cerchione et al., 2023).

In the upcoming section, we will delve into the third key attribute of Digital Nursing Quality Management - the digital ecosystem, and elaborate on its role in shaping the future of nursing practice.

#### Digital Ecosystem

The term “digital ecosystem” refers to the network of digital interactions among various entities interconnected through digital platforms, including individuals, organizations, services, and devices. Understanding and optimizing the digital ecosystem is critical in the context of Digital Nursing Quality Management.

##### (1) Holographic quantified digital environment

Digital ecosystems can also be focused into a myriad of smaller digital environments, where digital technologies are used to deliver, analyse, and integrate information and then process it algorithmically to form a controlled environmental space with a specific digital experience and feeling for the people who exist in it. On a digital level, it includes elements like the Internet, the Internet of Things, 5G, artificial intelligence, WeChat and their combined forms, such as electronic health records (EHRs), telemedicine platforms, wearable technology, and mobile health applications, while emphasizing a comprehensive understanding and mastery of the connections between these components (Ali et al., 2022). On a spatial level, it is an extension of the physical environment of human healthcare, and digitally transformed nursing quality data in any source and form is the core asset of this ecosystem, a holographic and comprehensive reflection of patient information with attributes and concerns different from those of the physical world (Hoyle, 2023).

##### (2) Evolution and Adaptability

The digital ecosystem combines technological and conceptual innovations, constantly evolving to meet reality’s needs (Kindermann et al., 2022). Stakeholders, including technology developers and organizational coordinators, adapt their activities with value propositions, fostering a virtuous cycle and evolutionary adaptation between operational capabilities, structure, and external environment (Pundziene et al., 2023).

By understanding and nurturing the digital ecosystem, Digital Nursing Quality Management can harness the full potential of digital technologies to enhance the quality of nursing care and patient outcomes. In the following sections, we will explore the antecedents and consequences of Digital Nursing Quality Management, further illuminating its role and implications in the healthcare context.

### Antecedents of digital nursing quality management

Antecedents are necessary precursors or conditions that lead to the emergence of a particular concept (Rodgers et al., 2018). Our literature review identified three principal antecedents associated with humans, organizations, and digital technology. These antecedents play significant roles in shaping the trajectory of digital nursing quality management, bearing a solid evolutionary link to the Donabedian model. However, this link is not a straightforward one-to-one correlation but rather an intricate interplay indicating the complex nature of this emerging field.

#### Human-related antecedents

The antecedents related to the human aspect predominantly involve nurses, patients, and their interpersonal process. The quantity and quality of nurses serve as crucial structural indicators of care quality and form an essential precondition for “personal involvement,” an inherent trait of quality management practices in nursing (Brommeyer et al., 2023). Nurses play a dual role, not just as the end-users of digital technology but as facilitators guiding patients and their caregivers in the configuration, utilization, and troubleshooting of digital health devices (Jedwab et al., 2023). Potential hindrances such as unfamiliarity with digital tools, deficiencies in digital skills, and apprehension regarding the learning curve of digital technology among nurses can diminish their propensity to adopt these technologies. This reduced inclination can impact the frequency and effectiveness of their usage of digital tools for monitoring and evaluating clinical care quality data, thereby hampering the successful implementation of digital nursing quality management practices (Dugstad et al., 2019, Gibson et al., 2009, Melkas, 2010). Furthermore, the human-centric focus of digital nursing quality management underscores the significance of patients as recipients of healthcare services. The concept of digital nursing quality management may remain elusive if patients resist adopting technology-driven solutions due to concerns regarding the usability of digital technologies or privacy infringements. This resistance is often called the “patient barrier” (Iyanna et al., 2022).

#### Organization-related antecedents

Organization-related antecedents encompass the structural and process indicators of the Donabedian model. They are captured through the four main categories of “organizational capability”: infrastructure, management, knowledge and learning, and collaboration (Gasco-Hernandez et al., 2022).

Firstly, the organization’s infrastructure significantly impacts how far quality management practice may advance. It is unrealistic to build a digital nursing quality management system in an environment without human resources, information, and assets (Dugstad et al., 2019, Lee et al., 2019). Secondly, as an evolving strategic approach, digital nursing quality management necessitates strong, persuasive leaders to create an organizational culture conducive to the implantation and diffusion of digital technologies to assist individuals in understanding organizational operations and providing them with shared values and belief patterns that regulate organizational behaviour, and to sort out functional requirements, data standards, technology management, vendors, and other complex issues (Jedwab et al., 2023, Kindermann et al., 2022). Thirdly, knowledge and learning are crucial antecedents for reflecting the “continuous improvement” aspects of quality management, which are frequently tightly linked to the capacity and thinking to recognize possible support and reconfigure and renew one’s resource base. Digital Nursing Quality Management requires innovative action by abandoning established, formal, and outdated organizational processes.

Finally, collaboration refers to the ability of organizations to successfully work together to implement plans or achieve goals, echoing the “interdependence and mutual benefit” of quality management (Gasco-Hernandez et al., 2022). This implies that a multi-centre consortium for digital nursing quality management is required, and they will play an important role in facilitating organizational cooperation, attracting funding, reducing duplication of effort, and distributing knowledge for technology deployment and adaptation (Cannavacciuolo et al., 2023, Dalenogare et al., 2023).

#### Digital technology-related antecedents

Digital technology and its successful adoption form the cornerstones for the inception and evolution of digital nursing quality management. They serve as a structural indicator of care quality and a stakeholder conduit to generate shared value. Firstly, according to the Unified Theory of Acceptance and Use of Technology(UTAUT), the acceptance and use of digital technologies by patients, physicians, nurses, and other stakeholders will emerge through an individual’s perceived usefulness of the technology, perceived availability, perceived physical condition, technological anxiety, and their own innovativeness (Edo et al., 2023). Patient’s willingness to accept digital technologies, nurses’ perceived usefulness and perceived ease of use of digital technologies, and their technical ability are all directly related to the successful implementation of digital nursing quality management practices (Choi and Tak, 2022, Wilson and Mooney, 2020, Wong et al., 2023). The better the device and the easier it is to operate, the more proactive nurses will be in analyzing the quality-of-care data, and the patients will be more accepting of the new technology (Au et al., 2019). Secondly, The propensity of nursing organizations to adopt technology is also influenced by peer evaluation of usage and acquisition costs, as well as whether digital technology is thoroughly utilized over time (Kateb et al., 2022). Finally, when information is quantified by digital technology and transformed into independent data as a critical component of evidence-based nursing decision-making and planning, data quality is crucial to demonstrate and evaluate the impact of an intervention, monitoring progress toward a goal, determining barriers to care, and influence public policy (Lindley et al., 2020).

### Consequences of digital nursing quality management

Consequences include incidents and events that occur as the results and outcomes of a concept (Tofthagen and Fagerstrøm, 2010). This study categorized the consequences into human-related consequences, organization-related consequences, and digital technology-related consequences.

#### Human-related consequences

Human-related consequences of digital nursing quality management predominantly manifest in patients and nurses. Patients benefit from digital nursing quality management because it assures that they receive timely, effective, and accurate treatment by facilitating information interchange and speedy decision-making within the ecosystem, increases patients’ personal care engagement, improves their perception of their health status and illness, and promotes self-efficacy (Farmer et al., 2017, Hong et al., 2020). Additionally, with digital nursing quality management, care activities and patient information are visible and traceable throughout the process, providing a legal basis for parties that need to assert their rights (He, 2022).

Nurses achieve continuous, dynamic assessment and management of patients with the support of information technology, real-time, comprehensive patient information, and more targeted decision-making, reducing the burden and improving efficiency (Arakawa et al., 2018, Au et al., 2019, Dugstad et al., 2019, Roeder, 2009). Additionally, nurses can leverage digital tools to provide timely feedback, address queries, and offer emotional support, ultimately promoting improved health outcomes and patient satisfaction. In contrast, digital performance expression and public administration of evaluation results boost nurse motivation (Aapro et al., 2020).

#### Organization-related consequences

The organization has an essential role in digital nursing quality management. On the one hand, digital nursing quality management enhances communication between nursing departments and units, enables more accurate staff allocation and staffing within each organization, improves management efficiency, reduces overstaffing or overload, and drives organizations from “empirical and institutional” management to “data-based” management (Back et al., 2022, Bail et al., 2023). Simultaneously, multiple single digital ecologies are connected to form a wider-coverage information and decision-making sharing network, facilitating inter-organizational collaboration and giving a larger sample of data and practice references that drive management model optimization (Cherbib et al., 2021, Philpot et al., 2023). On the other hand, the ideals of “digital humanism” have an environment in which they might take root and thrive. Organizations begin to establish digital strategies that consider the growth of digital value activities in a humanistic care environment, inspire their members’ behavior, lead them to adopt digital technology as a source of value and share and produce information. It is evident that the development of digital technology has had a profound impact on healthcare professionals and organisations. The large amount of healthcare data generated via technology-based assistance, as well as the appropriate use of such information and data, promotes the sharing, monitoring, tracking, and analysis of a patient’s heath by both the patient and the healthcare provider (Varshney, 2007). Yet, issues of data security and data privacy exposed during the operation of the digital ecosystem should be given equal consideration. There are several concerns with the exchange of medical information, and medical data can be readily tampered with or lost during the transmission process. Furthermore, hospitals hold the majority of medical data, and the data is likely to be sent and used without their permission, posing security problems and causing conflicts between healthcare providers and patients.

#### Digital technology-related consequences

Investigation into the digital technology-related consequences showed that new technologies do not always imply improved patient health outcomes and quality management. When digital technology is utilized clinically without being tested in stages or as a “complementary” tool rather than a “replacement” product, it can lead to under- and overuse of care (low-value care) and negative usage evaluations, thus providing a realistic basis for a new round of technology turnover that can be tracked (Laine et al., 2023, O’Reilly-Jacob et al., 2021). It is also important to note that digital technology, as a new link between nurse and patient, simplifies the nursing process while simultaneously reducing face-to-face interaction. Promoting digital technology’s humanization by drawing on nursing’s innate humanist connotations is a critical issue to consider at this time (Ali et al., 2022, Oberg et al., 2018).

In summary, digital nursing quality management is a new type of ecosystem with a high degree of synergy and co-sustainability of multiple elements, with tremendous potential to improve patient health outcomes, improve the quality and efficiency of healthcare services, and optimize the humanistic content of the digital environment. More notably, the digital nursing quality management system transforms “value co-creation” from vision to reality. According to “Service-Dominant Logic” (S-D Logic), value is co-created in healthcare through the interaction of patients and providers: the former has information, while the latter can provide education as well as management and analysis of the patients’ information (Balta et al., 2021, Ebneter et al., 2022, Mackow et al., 2023). When digital technology is integrated into the healthcare delivery system and becomes an important medium, patients, providers, professionals, technology, and information all become important resource components of the system, and the empowerment of their interactions opens up possibilities of action for further optimization. Value, thus, for healthcare (and our research) does not only mean business value but also societal value that leads to long-term healthcare and societal development.

### Related term

Reviewing terms related to the concept can facilitate more specific exploration and analysis, especially for analysing any differences from the central concept (Rodgers et al., 2018). In the literature, we found that the terms “Intelligent Health Information Service” and “Full Life-Cycle Health Management” are closely related to digital nursing quality management.

Intelligent health information service evolved from digital health information service, which is an expression of the interdisciplinary field of intelligent health (Ma and Zhou, 2018). The use of modern information technology and intelligent technology to improve the traditional medical and health management model is at the core of intelligent health, while intelligent health information service emphasizes wisdom, utilizing cloud computing, artificial intelligence, the Internet of Things, sensors, and other advanced technologies to achieve personalized, convenient, networked, and intelligent health information services (Schaefer and Atreya, 2020). Because the intelligent health information service is still in its early stages of development, academics have yet to establish a rigorous and consistent conceptual framework. Full life-cycle health management refers to the ongoing and tailored provision of health management services to residents based on the human life cycle and in response to the features of health demands at various phases (Wilson et al., 2021). There are still issues in the current medical and health service system, such as insufficient information exchange between subjects, a low degree of information integration, and inefficient information consumption (Green et al., 2023, Ow Yong et al., 2023, Wu et al., 2023). The public’s access to lifecycle health management services is hampered by the lag in healthcare delivery. To resolve this dilemma, it is necessary to realize the digital transformation of the healthcare service system through technological coupling, as well as to establish a digital nursing quality management system that is interconnected, intelligently perceived, and interacts with real and virtual reality, so that each subject can collaborate appropriately to help realize the full life-cycle healthcare services.

### Model Case

The Rodgers’ evolutionary method encourages identifying a model case that utilizes all the necessary attributes depicted in the literature analysed. A model case is designed to showcase an example of the concept with all its attributes in place.

The Teays Valley Centre, located in West Virginia, USA, is a subsidiary of Genesis. It is a skilled nursing facility with 128 certified beds offering various services, including wound care ***(organization-related antecedents)***. In 2015, the facility decided to implement an electronic wound care documentation and management system through the findings of a quality improvement study ***(organization-related antecedents)***, and in 2017 added the position of Skin Integrity Coordinator (SIC) as a core leader with primary responsibility for coordinating all activities related to wound care management ***(individual-related antecedents, organization-related antecedents)***. The facility uses a skin and wound app to replace the traditional ruler-based measurement and mapping methods for focal care ***(digital technology-related antecedents)***. Information from each patient consultation is stored encrypted on a cloud-based server. Wounds are monitored and tracked via a computer-based skin and wound dashboard that also allows real-time analytics and facilitates the secure, easy, and bulk distribution of wound information (e.g., wound photos) among interdisciplinary teams (IDT) members ***(human-machine collaboration, digital Ecosystem)***. If complicated wounds are identified that cannot be treated appropriately and managed inside the institution, the patient and their treatment information are forwarded to a specialized wound care centre for treatment simultaneously ***(digital Ecosystem)***. After a period of application, the facility found that the implementation of a digital skin and wound management system made it possible for nurse practitioners to thoroughly investigate all wounds in the facility ***(interactive empowerment, individual-related consequences)***, while the digital photos effectively compensated for the manual written descriptions that did not accurately convey the status of the wounds and were more conducive to improve treatment decisions and overall wound management. It provided the patient or institution with the legal requirements for litigation evidence ***(individual-related consequences, organization-related consequences)***. In addition, the digitization of wound information and data-driven risk identification significantly reduces the burden and workload of nurses, ensures accurate wound classification, and increases the transparency of the wound management process ***(individual-related consequences, organization-related consequences)***.

### Definition of Digital Nursing Quality Management

Digital Nursing Quality Management is a strategic management approach that systematically utilizes evolving digital technology to pursue optimal nursing quality, guided by the timeless principle of humanistic care. The model of Digital Nursing Quality Management is shown in Fig. 2.

## Discussion

*“I will remember that there is an art to medicine as well as science and that warmth, sympathy, and understanding may outweigh the surgeon’s knife, the chemist’s drug, or the programmer’s algorithm.”*

As this revised Hippocratic Oath (Meskó and Spiegel, 2022) poignantly underscores, the humanistic foundations of care remain paramount in the rapid evolution of digital healthcare technology. Within this dynamic landscape, we now focus on the new concept of Digital Nursing Quality Management. Through our analysis, we strive to honor this timeless principle of humanistic care while simultaneously exploring the transformative potential of digital technologies in reshaping the nursing profession.

In this study, we determined the three defining attributes of Digital Nursing Quality Management (DNQM) and clarified the antecedents, consequences, and a model case in a real-world setting related to DNQM. Based on the results of the conceptual analysis, the operational definition of “digital nursing quality management” is “ a strategic management approach that systematically utilizes evolving digital technology to pursue optimal nursing quality, guided by the timeless principle of humanistic care.” We have embodied in detail in our conceptual model of digital nursing quality management how people, organizations, and digital technologies can empower each other and form a patient-centred value co-creation system. In an era of increasing convergence between digital technologies and the healthcare field, identifying the subjects involved, the resources required, and potential courses of action for a strategic approach to digital nursing quality management can maximize the healthcare potential of emerging digital technologies and is critical in terms of quality and efficiency gains in nursing, as well as improvements in patients’ experience of care and health outcomes.

Furthermore, this analysis provides an opportunity for future research studies and practice. Few studies, for example, have focused on how inadequately mature digital technologies in clinical practice impact nurse quality management behaviors and results, and the truth is that the technologies used in real-world healthcare settings are not totally responsive or completely appreciated(Webster, 2020, Whitmore et al., 2020). Equally of interest is whether the human touch, which has been valued in traditional face-to-face interactions, will be undermined by the incorporation of technology, even as the emergence of emerging digital media between caregivers and patients allows for improved quality and efficiency in care delivery. There is an undeniable need in the rapidly evolving healthcare sector to enhance nursing management by utilising the complementary strengths of humans and machines, the former with their unique capabilities of critical thinking, empathy and contextual understanding, and the latter with their strengths of data processing, pattern recognition and operational efficiency (Ali et al., 2022). Therefore, how to narrow the distance between user needs and actual technology, how to integrate humanist overtones of care into each iteration of digital health technology, and how further to deepen human-machine collaboration into a harmonious human-machine symbiosis are all crucial issues to consider at the moment.

### Limitations

In acknowledging the boundaries of our concept analysis on digital nursing quality management, we recognize several limitations that should be considered in interpreting our findings and used to inform future research directions.

Firstly, the concept of digital nursing quality management is subject to constant evolution due to continuous innovations in digital technologies. Therefore, the attributes identified in this study may evolve as new technologies emerge and reshape the landscape of nursing practice.

Secondly, the scope of our literature review was limited to specific databases focusing on English and Chinese-language publications. This limitation could potentially restrict the diversity of digital nursing quality management perspectives, excluding critical insights from other linguistic and cultural contexts. Future research could incorporate a broader range of databases and languages to enhance the comprehensiveness of their review.

Additionally, while our study emphasized scholarly literature, it excluded grey literature. Although this approach maintained academic rigor, it might have overlooked practical insights from sources like blogs, white papers, or policy documents. In particular, blogs authored by influential figures in the emerging digital nursing quality management field may provide valuable, up-to-the-minute insights that complement academic studies. Future research should consider including these resources to augment their understanding of the concept.

Lastly, our analysis emphasized specific digital nursing quality management attributes, which might have excluded certain cultural and context-related aspects. While this focus provided an in-depth examination of critical attributes, it could limit the applicability of our findings across different healthcare settings. Future studies could encompass these aspects to improve the transferability of their findings.

In conclusion, while we have conducted this concept analysis with meticulous care, we must consider these limitations when interpreting our findings. Future digital nursing quality management researchers can utilize these limitations as guideposts, enabling a more comprehensive understanding of this evolving concept.

## Conclusions

This concept analysis has comprehensively elucidated digital nursing quality management by applying Rodgers’ evolutionary method. Our exploration started with examining the principal attributes, including human-machine collation, interactive empowerment, and digital ecosystems, identifying antecedents, and analysing the potential consequences. The Teays Valley Centre model case further illuminated the concept’s practical implications, highlighting the transformative potential of digital technologies in the sphere of nursing care. The findings suggest that digital nursing quality management is not a mere amalgamation of separate components but a dynamic ecosystem shaped by individual, organizational, and technological factors.

## Data Availability

All data produced in the present work are contained in the manuscript.

